# What successful adaptation reveals about capability and dependence in healthcare: A Brittle Threshold perspective

**DOI:** 10.64898/2026.10.28.26366747

**Authors:** Sayra Cristancho, Crystal Gaudet

## Abstract

Healthcare teams routinely adapt to changing demands by redistributing tasks, stretching roles, and reorganizing coordination. Although these adjustments can sustain performance, their immediate success provides limited insight into how they affect the conditions supporting subsequent adaptation. We examined how successful adaptation reorganizes collective work and what these changes might reveal about emerging limits to adaptability.

Guided by Brittle Threshold as a sensitizing concept, we conducted a secondary qualitative analysis of 34 anonymized interviews from four studies involving hospital teams during COVID-19, military surgical resuscitation teams, rural emergency care teams, and a community-led mass vaccination initiative. The datasets provided accounts of adaptation under varied demands and constraints.

Across settings, adaptation changed collective work through four connected domains: work redistribution, coordination reconfiguration, responsibility concentration, and human requirements. Similar adaptations were sustained under different conditions. Some were accompanied by shared learning, expanded skills, and alternative sources of support, while others maintained performance through continued dependence on particular individuals, restricted access, or sustained personal effort. These patterns could coexist within the same setting. Examining the relationships among the four domains revealed differences in how collective performance was organized and supported that were not apparent from successful task completion alone.

The findings offer an empirically grounded approach to examining potential Brittle Threshold signals in everyday healthcare work. Rather than identifying particular adaptive behaviours as warning signs, they direct attention to changing configurations of responsibility, coordination, support, and human effort. This perspective can inform investigations of how healthcare teams sustain adaptability as demands change.

## Introduction

Healthcare teams routinely adapt when demands exceed what usual ways of working can accommodate. During crises and under resource constraints, people redistribute tasks, work across role boundaries, improvise solutions, and change how they coordinate to maintain care. Research has documented how these responses vary with team structures, membership, and the circumstances in which work takes place (Sanford et al., 2025). Such adaptations can be essential to successful performance, yet their immediate success provides an incomplete account of what they accomplish. Workarounds can resolve frontline problems while concealing resource shortages and reducing pressure to address them (Morrison, 2015). Healthcare delivery can also depend on coordination, judgment, and additional work that remain poorly recognized within formal accounts of service provision (Barnard et al., 2024). Understanding successful adaptation therefore requires attention to how performance is maintained and how adaptation changes the way people organize and carry out work together.

Maintaining performance during a particular demand does not establish that a team remains able to adapt when demands change again. Resilience engineering distinguishes successful responses to disturbances from sustained adaptability across changing conditions (Woods, 2015, 2018). Adaptive capacity is finite, and the relationships through which people and units support one another can extend or constrain their ability to respond. Successful responses may also generate further demands, changing what subsequent adaptation requires (Woods, 2018). Team adaptation scholarship similarly conceptualizes adaptation as a recursive process, in which adjustments can modify cognitive, social, and organizational resources relevant to subsequent activity (Burke et al., 2006; Maynard et al., 2015). Together, these perspectives establish why the consequences of adaptation matter beyond the immediate demands it addresses.

Brittle Threshold names the zone in which adaptation continues to sustain performance while becoming increasingly fragile (Cristancho, 2026). The concept brings attention to a difficulty in recognizing adaptive limits. When people continue to meet demands, their success can be taken as reassurance that they can keep doing so. Yet the work required to achieve that success may be becoming harder to sustain. Brittle Threshold keeps this possibility open for examination, prompting questions about what people are drawing upon to keep work going and whether they can continue to do so as demands change. For this study, the concept raises an empirical question about how that fragility might become recognizable in everyday work before performance deteriorates.

There is already evidence that changes in everyday work can provide information about capacity strain. Womack et al. (2019) identified workplace activity changes and adaptive strategies that experienced nurses recognized as cues of imbalance between patient demand and available capacity. Hionis et al. (2026) examined coordination patterns as markers of system brittleness, connecting those patterns to the pressures and adaptations through which work was accomplished. However, role flexibility, mutual support, and changing coordination can also be important features of effective collective performance (Lingard, 2016). Recognizing an adjustment therefore does not, by itself, establish what it signals. The challenge is to understand whether changes in adaptive work warrant concern even as that work continues to succeed.

Guided by Brittle Threshold as a sensitizing concept, this study sought to identify and characterize changes in adaptive work that might signal increasing fragility while performance remained successful. The aim was to develop an empirically grounded account of potential signals to inform prospective studies of changing conditions of adaptability.

## Methods

### Study design and conceptual orientation

We undertook a secondary qualitative analysis of four existing datasets examining how people adapted to crisis conditions and changing demands. The datasets included hospital teams working during COVID-19 (ATA), military surgical resuscitation teams (MSRT), a community-led mass vaccination initiative (CoVax), and rural emergency care teams (RP). Together, these studies provided accounts of how healthcare and community teams maintained work under different demands and constraints.

Brittle Threshold informed the analysis as a sensitizing concept (Cristancho, 2026). The concept directs attention to the zone in which adaptation remains effective while the conditions supporting it become increasingly fragile. Our initial purpose was to explore whether participants’ accounts contained perceptible changes in adaptive work that might serve as potential signals of this condition.

We used *signals* provisionally to refer to changes in adaptive work that warranted further examination. We did not treat them as validated indicators or assume that they predicted subsequent failure.

### Data sources and selection

The secondary analysis drew on 34 anonymized interview transcripts from four previously conducted studies (ATA, n=5; MSRT, n =5; RP, n=12; CoVax, n=12).

- Hospital teams working during COVID-19 (ATA): Using constructivist grounded theory (CGT), this study explored the strategies healthcare teams used to adapt during the COVID-19 pandemic. Semi-structured interviews were conducted with 20 healthcare workers from hospitals across Ontario, Canada (Cristancho et al., 2022). Four transcripts from this dataset were included in the secondary analysis.
- Military surgical resuscitation teams (MSRT): This qualitative descriptive study examined the functions and foundational underpinnings of how military surgical resuscitation teams approach crises. Fifteen semi-structured interviews were conducted with members of surgical resuscitation teams within the United States Special Operations Command (Eckert et al., 2024). Five transcripts were included in the secondary analysis.
- Community-led mass vaccination initiative (CoVax): The CoVax study explored a collaborative, community-driven initiative to establish a COVID-19 vaccination clinic in Cobourg, Ontario, Canada (Gaudet et al., 2024). Semi-structured interviews were conducted between September 2022 and July 2023 with 34 individuals involved in coordinating and operating the clinic, including community volunteers, hospital and public health unit staff, members of local community organizations and businesses, and municipal employees. Twelve transcripts were included in the secondary analysis.
- Rural emergency care teams (RP): This CGT study explored plasticity in rural healthcare practice through semi-structured interviews with physicians and nurses working in two emergency departments in Ontario, Canada (Gaudet et al., 2026). The original study included 19 interviews conducted between January and November 2024. Twelve transcripts were included in the secondary analysis.

The datasets were selected because they captured adaptation across varied settings, team compositions, and demands, providing an opportunity to examine potential brittle threshold signals across distinct contexts. This variation allowed us to consider how changes in adaptive work might be expressed under different conditions, while attending to both shared patterns and context-specific features.

A subset of transcripts from each dataset was selected for inclusion in the secondary analysis. Where possible, selection aimed to capture variation in participant roles within and across datasets, allowing us to examine potential brittle threshold signals across different roles and experiences.

### Analytical process

Our analysis began by revisiting participants’ accounts with attention to how teams adapted while continuing to deliver care or services. We were particularly interested in changes that might reveal growing strain or emerging limits to adaptation.

SC and CG independently reviewed and coded one transcript from each study setting in full. They then met to discuss their coding and identify accounts in which adaptive work appeared to involve signs of strain, changing demands, or potential limits to teams’ capacity to sustain performance. This initial review supported a shared understanding of the analytic focus while allowing the researchers to approach the accounts from their respective perspectives.

CG subsequently read and coded the remaining transcripts, meeting with SC every three to five interviews to discuss emerging codes, categories, and patterns across the data. These discussions supported the ongoing refinement of the analysis and consideration of how emerging interpretations related to the study’s focus on brittle thresholds.

The data were organized and coded using Quirkos. Rather than applying predetermined indicators, we remained attentive to patterns emerging through engagement with participants’ accounts. Initial coding focused on how teams adapted to sustained demands and constraints, including emotional and moral strain associated with sustained adaptation, the normalization of improvisation under constraint, accountability drift toward the frontline, informal bridging of organizational gaps, persistent resource scarcity and compensatory workarounds, and role saturation.

As the analysis developed, we recognized that changes in adaptive work could reflect the development of collective capabilities as well as continued reliance on particular people, relationships, and additional effort. This broadened our examination from possible signs of fragility to how successful adaptation reorganized collective work and what those changes revealed about the conditions sustaining performance.

We subsequently examined accounts across the datasets to understand how changes in collective work were related. Through discussion and comparison of the emerging codes, we collaboratively refined the final codes and developed four domains of analysis: work redistribution, coordination reconfiguration, responsibility concentration, and human requirements. These domains brought together related patterns in participants’ accounts while providing a way to examine different aspects of adaptive work across contexts.

Comparisons across the datasets also highlighted differences in how participants understood and experienced adaptation. Although our initial analysis focused on potential signals of strain, participants’ accounts did not always frame adaptation in these terms. For example, in CoVax, participants often described the initiative as a success. Attending to these accounts prompted us to consider how adaptive work could support successful outcomes while also examining the conditions and efforts on which those outcomes depended, including how similar adaptive responses could have different implications across contexts.

### Researcher positioning

SC was the principal investigator on all four original studies, providing familiarity with their research contexts, aims, and data. CG was involved in the original CoVax and rural plasticity studies but was not involved in the original ATA or MSRT studies. The researchers brought different levels of familiarity with the datasets to the secondary analysis, which informed their discussions of emerging interpretations.

### Ethics

The four original studies received approval from Western University’s Health Sciences Research Ethics Board (Review Reference Nos. ATA: 2019-113589-23481; MSRT: 2024-124712-91231; RP: 2023-121975-75949; and CoVax: 2022-121261-69644). The present secondary analysis was conducted using previously collected, anonymized interview transcripts. No additional data were collected from participants.

## Results

### 1. Adaptation reorganized collective work

Across four settings, people responded to changing demands by stretching roles, redistributing tasks, improvising, crossing professional boundaries, drawing on relationships, and changing how they coordinated. Participants described these responses as enabling care or services to continue. They also changed how people organized and carried out the work together.

We use ‘collective work’ to refer to the work through which people jointly accomplish what a situation requires. This directs attention to how work is organized among people, not only to individual actions. In rural emergency departments, nurses described absorbing registration, laboratory, IT, maintenance, and other functions while continuing to provide patient care. As one nurse put it, they were *“usually running the emerg and taking care of patients at the same time”* (RP Nur 1). Work that would ordinarily be carried out elsewhere had moved to those remaining on site. A physician likewise described the additional work taken on when usual services were limited: *“you end up calling family meetings. I’ve done like neurocognitive testing… which is not really emergency department material, but that’s what you’re doing… We have limited physio… So, we do a lot of walk testing for people, like we do a lot of gait testing”* (RP Phy 1). These changes involved more than the addition of tasks. They altered who performed the work, how it was coordinated, and who remained responsible for its completion.

In comparing these accounts, four connected aspects of collective work became salient: work redistribution (or where the work went); coordination reconfiguration (or how people organized and enabled it); responsibility concentration (or where responsibility accumulated or could be shared); and human requirements (or what maintaining the work asked of people). The first three draw attention to how collective work was organized. Human requirements make visible the time, expertise, attention, availability, and practical or emotional effort needed to sustain that work. These were not four separate types of adaptation. Their importance lay in what their relationships revealed about how performance was being maintained.

### 2. Similar adaptations could leave the collective differently positioned for what came next

Working beyond usual role boundaries did not always have the same implications. In military surgical resuscitation teams (MSRT), role flexibility was deliberately cultivated through cross-training and exercises. Team members were sometimes placed in roles other than their own during exercises to *“actively try to enable those skills”* (MSRT P10). The intention was to make different people better prepared to undertake work that might be needed when demands changed.

Participants also described using after-action reports to document lessons from exercises and inform subsequent training and scenarios. One described revisiting changes in subsequent exercises to ask, *“did that work or did it make it worse, what was better?”* (MSRT P2). Participants also described practical backup arrangements. A surgeon explained that the team tried to *“have a backup ready at short notice to take over,”* with two MSRT surgeons at the same institution covering one another when possible (MSRT P5). Role flexibility here was supported by opportunities to learn, share knowledge, and arrange substitutes. These accounts demonstrated deliberate preparation and reported changes to training and coverage.

In the community vaccination response (CoVax), participants described developing a system for work they had not previously done together. A small group worked out patient flow, vaccine movement, information requirements, and related logistics. As experience accumulated, the system *“morphed and got better and better”* (CoVax Par 25). Participants described revising procedures between shifts and learning how colleagues worked. One volunteer explained that they *“ended up all learning how to work together, respecting and trusting each other just because we had worked together so long”* (CoVax Par 5). The team also, *“ended up setting up weekly meetings to do debriefs and discuss issues”* (CoVax Par 31). Participants described learning and ways of coordinating that could be carried into subsequent clinics.

In both MSRT and CoVax, participants described adaptations accompanied by deliberate efforts to develop skills, share learning, and organize support. The observable difference was not role flexibility alone. The significance of that flexibility depended on whether others could participate, learn, or step in when needed.

### 3. Other adaptations maintained performance through continued dependence

In the rural data, people also stretched roles and redistributed tasks, but some work remained difficult to pass to others. A nurse described administrative tasks associated with transferring a patient that could not be delegated to colleagues from another ward. They might have helped, yet calling EMS for a transfer was something *“only we do and only we have a login for*.*”* As the nurse explained, *“delegating an administrative task would be really helpful, but they don’t actually have a login for it. So, you can’t do that”* (RP Nur 1). Staff could be available in the building without being able to assume the work. Technical permissions and familiarity with procedures shaped who could help and where responsibility remained.

The relationship between unavailable services, concentrated responsibility, and human requirements was also visible when several demands collided. Another nurse described a food-service failure while caring for ten patients in the emergency department, including one receiving a norepinephrine infusion. After attempts to resolve the problem through management, the nurse was told, *“you’re going to have to make dinner*.*”* The nurse responded: *“I’ve got 10 people in my Emerg, someone on a norepi drip. I don’t have time to go make dinner for six patients down the hall”* (RP Nur 4). Elsewhere in the same account, the participant described making grilled-cheese sandwiches with the supplies available. The immediate food need was met, but doing so required clinical staff to absorb food-service work while already responsible for patients needing care. In this episode, the solution relied on the nurse rather than an alternative food-service arrangement.

These examples showed why the work’s movement matters. People could keep tasks from going undone, but the ability to maintain service remained tied to particular staff, access arrangements, and additional effort. This pattern did not characterize all rural adaptation. Rather, it revealed one way that successful task completion could remain dependent on the people already carrying the work.

### 4. Capacity-building and continued dependence could coexist within the same setting

The changes in collective work described in the preceding sections did not divide neatly by setting. CoVax, for example, developed processes for shared learning while some activities continued to require considerable individual effort. One participant described the practice of recording what could be improved after each vaccination shift and implementing changes at the next: *“At the end of each shift, [we] wrote down what they would do differently, how you would do it differently, what you liked about it, what didn’t work, and implemented it right away”* (CoVax Par 29). Another described debriefs and morning meetings that helped maintain communication as requirements changed rapidly, while acknowledging that the work was *“overwhelming for everybody”* (CoVax Par 31). Participants reported both deliberate refinements and substantial demands on those coordinating the work.

At the same time, mobile vaccination work required prolonged effort under difficult conditions. A participant who had helped develop the initial vaccination processes subsequently described vaccinating people outdoors without running water, bathrooms, or sufficient staffing to take breaks.

After repetitive work in the heat, *“my brain just kind of went off”*; the participant recalled asking a person, *“Have I given you the needle yet?”* (CoVax Par 25). Another participant described working “*ten-hour days. Did it mean that we had our 15-minute breaks every two hours or a one-hour lunch? No, not necessarily*.*”* (CoVax Par 30). These accounts showed what continued delivery was requiring of those performing it.

The mobile team reported delivering 2,000 vaccinations over a couple of weeks. As one participant recalled, *“the health unit would let us do a few more with their vaccine because they saw we could do it and we were getting the job done”* (CoVax Par 25). The team’s success opened opportunities to extend vaccination services, building on a way of working that required considerable personal effort.

CoVax thus developed learning routines that made some activities easier to organize alongside work sustained through extended availability and effort. Capability-building and concentrated human requirements could coexist within one initiative, even where participants described operational success. What mattered analytically was how collective work was organized and supported across these activities.

### 5. The four domains acquired meaning through their relationships

Examining the domains together showed how changes in collective work depended on the access, skills, and coordination available to support them. In the rural transfer account, the administrative task remained with the emergency nurse because colleagues lacked system permissions. Other rural accounts showed how tasks could be shared more easily. As registered practical nurses became more comfortable working within their scope of practice, one participant explained that *“they’ll just go straight to the doc”* instead of relying on the RN to relay information (RP Nur 5). Another described how personal support workers helped clinicians contact respiratory therapists while attending to a patient, explaining that *“they can make the call for us and put us on speaker phone”* (RP Nur 2). Across these examples, access to systems and communication with colleagues shaped how tasks and coordination could be shared.

The significance of responsibility concentration likewise depended on available alternatives. MSRT participants described planning for short-notice coverage when clinicians deployed; by contrast, an ATA physician recalled a period when residents became unavailable and *“there wasn’t the redundancy in the system”* (ATA-C-002). The participant feared that if another clinician became ill there might be nobody to step in. The same physician later reported, *“well, now, there is a backup schedule,”* which had been introduced toward the end of the third week. Across these accounts, the availability of backup shaped how responsibility could be shared when clinicians were absent.

Supporting MSRT deployments required coordination with the civilian departments where military clinicians also worked. One participant explained that when clinicians were called to deploy, *“they create holes in their schedule”* that civilian departments had to fill (MSRT P2). Before one deployment, the military team they were supporting agreed to shorten the planned pre-deployment training, explaining that *“we can condense the training from five days to two”* (MSRT P2). This adjustment helped the MSRT prepare for deployment while reducing disruption to civilian staffing.

Human requirements also varied with the support available for different activities. In CoVax, participants described debriefs and shared learning that helped coordinate clinic work, alongside mobile vaccination where limited staffing meant long shifts without regular breaks.

Across the cases, examining the four domains together revealed how teams could become better equipped to share some work while continuing to depend heavily on particular people for other tasks. New ways to communicate, learn, and provide backup expanded support for some activities, while restricted access, staffing gaps, and sustained personal effort kept other tasks concentrated. These conditions could coexist across activities within the same setting, and participants described how they changed as teams developed new ways of working. These differences were visible even as teams continued to deliver care and services.

## Discussion

Our findings show why the significance of successful adaptation cannot be determined from the adaptive response alone. Across four settings, work was redistributed, roles stretched, and coordination changed while care or services continued. Yet these adjustments were sustained through different arrangements of access, support, responsibility, and human effort. Team adaptation scholarship has long established the importance of role changes, collective processes, and emergent coordination (Burke et al., 2006; Maynard et al., 2015; Sanford et al., 2025). Our findings contribute an account of how four connected domains – work redistribution, coordination reconfiguration, responsibility concentration, and human requirements – make the organization of successful adaptive work empirically examinable. Reading the domains together helps explain why outwardly similar adjustments can reveal different conditions for maintaining collective performance.

Those conditions matter beyond the immediate response. Woods (2015, 2018) distinguishes successful adaptation to a disturbance from sustained adaptability as demands change. Capabilities are finite and relational, therefore, what one person or unit can accomplish depends in part on the support available elsewhere. Wikström and Gadolin (2026) show how healthcare services may continue through sustaining work that becomes embedded in shared routines and infrastructure, or through compensatory labour continually supplied by particular individuals. Our analysis specifies how different forms of support and dependence become recognizable in the organization of adaptive work. Redistributed tasks, for example, have different implications when people also acquire the access and skills to undertake them than when responsibility remains with those who already carry the work. Human requirements bring into view the continuing effort and availability on which these arrangements depend, without assuming that such effort is inherently problematic.

An adaptation can also support work locally while shifting requirements across people, services, or organizational boundaries. Woods’s (2018) account of interdependent adaptive units explains why extending one unit’s capability may depend on accommodations elsewhere. Our MSRT material offers a concrete illustration, where maintaining readiness for deployment could require civilian departments to reorganize staffing, and the timing of training was negotiated to limit disruption to those departments. Other accounts described organizational routines being revised after a gap in support became apparent. These observations caution against treating a team’s adaptive capability as a self-contained attribute or interpreting dependence as a fixed property. Whether a particular configuration of collective work remains workable as circumstances change may depend on the support and alternatives available. Understanding how these conditions develop across successive demands will require prospective research.

The findings also refine what we mean by a potential Brittle Threshold signal (Cristancho, 2026). Previous research has identified changes in everyday activity as cues of capacity strain (Womack et al., 2019) and coordination patterns as markers of system stretch (Hionis et al., 2026). We do not propose another checklist of adaptive behaviours. Each domain is ambiguous when viewed alone. For instance, responsibility may reasonably concentrate around expertise, informal coordination may be effective, and substantial effort can be part of competent collective work (Lingard, 2016). What warrants attention is how these elements come together. Work may become more shareable through developing capability while certain tasks continue to rely on a small number of people; new coordination arrangements may extend support while requiring continuous availability. In this account, a potential signal is a perceptible change in how collective work is organized that opens inquiry into what is sustaining performance. It does not diagnose proximity to a threshold or predict failure.

These distinctions have implications for the social organization and management of healthcare work. Operational continuity can remain visible while the coordination, judgment, and additional labour needed to sustain it are poorly recognized. Morrison (2015) shows how successful workarounds can conceal shortages; Barnard et al. (2024) document the hidden work and professional commitments sustaining everyday care; and Brisley et al. (2025) show how organizational improvement can concentrate additional work among particular staff. Our findings suggest that decisions about staffing, service provision, technical permissions, and coordination would benefit from examining where responsibilities are being carried and what makes their redistribution possible. The issue is also one of perspective. People undertaking the work may recognize dependencies and human requirements that are less apparent in organizational accounts of successful performance. Our data do not show that managers reduced resources because staff continued to cope, but they illustrate why continued functioning alone is insufficient evidence of adequate support.

This secondary analysis offers a retrospective view of adaptation. The original interviews explored how people responded to demanding situations. Accounts of continued service delivery or completed targets cannot establish the safety or quality of every action, and training exercises provide different evidence from clinical deployments. Our analysis shows how collective work changed and what people relied on to keep it going. It cannot tell us how close teams were to an adaptive limit or how these ways of working would hold up under new demands. Prospective research could follow the four domains over time to examine how teams develop shared capability, where work continues to depend on particular people and their effort, and how these patterns change together. This would help us understand what successful adaptation leaves teams able to draw on when demands change again.

### AI Disclosure

During the preparation of this manuscript, the authors used ChatGPT to support the development of the manuscript’s narrative and to refine the clarity and effectiveness of the writing. The authors reviewed and edited the AI-assisted content and take full responsibility for the final manuscript.

## Data Availability

All data produced in the present study are available upon reasonable request to the authors

